# Genetic risk scores of disease and mortality capture differences in longevity, economic behavior, and insurance outcomes

**DOI:** 10.1101/2020.03.30.20047290

**Authors:** Richard Karlsson Linnér, Philipp D. Koellinger

**Affiliations:** School of Business and Economics, Vrije Universiteit Amsterdam, De Boelelaan 1105, 1081HV Amsterdam, The Netherlands

**Author notes:** &.

## Abstract

Widespread genetic testing for diseases may cause adverse selection, escalating premiums, or discrimination in various insurance markets. Here, without systematically informing study participants of their genetic predisposition, we estimate to what extent genetic data are informative about differences in longevity, health expectations, and economic behavior. We compute measures of genetic liability (polygenic scores) for 27 common diseases and mortality risks in 9,272 participants of the Health and Retirement Study (HRS). Survival analysis suggests that the highest decile of cumulative genetic risk can distinguish a median lifespan up to 4.5 years shorter, a difference that is similar to or larger than that distinguished by conventional actuarial risk factors, including sex. Furthermore, greater genetic liability is associated with less long-term care insurance, among other economic behaviors. We conclude that the rapid developments in genetic epidemiology pose new challenges for regulating consumer genetics and insurance markets, requiring urgent attention from policymakers.

## 1 Introduction

The precision of genetic tests for common diseases is rapidly increasing. While the prognostic accuracy of these tests is still limited, it will become substantial in the near future (Zeggini et al. 2019). Recent studies find that cumulative measures of genetic liability, which are called *polygenic scores*, are comparable in precision to other, established clinical risk factors (Abraham et al. 2019; Khera et al. 2019; Torkamani, Wineinger, and Topol 2018). Polygenic scores have quickly become a routine component of genetic health reports offered by companies in the exponentially growing market of consumer genetics, with several millions of customers worldwide (Khan and Mittelman 2018). Many customers report a strong motivation to explore their DNA for health information (Nelson, Bowen, and Fullerton 2019; Wang et al. 2018). Thus, genetic testing is becoming an affordable and widely advertised service, and people can now purchase more or less noisy estimates of their genetic liability early in life, many years before any signs or symptoms of disease emerge (Khera et al. 2019). This study estimates to what extent polygenic scores for a range of common diseases and mortality risks from various medical domains can capture differences in longevity, subjective life expectancy, self-rated health, and insurance purchases, and we discuss the implications of our findings.

In summary, we generated polygenic scores for 13 common medical conditions related to mortality (including Alzheimer’s disease, atrial fibrillation, and type 2 diabetes) and 14 mortality risk factors (including blood pressure, cholesterol, and smoking) by leveraging results from the largest genetic studies thus far made publicly available to the research community. In a series of survival analyses in 9,272 Health and Retirement Study (HRS) respondents, we found that the polygenic scores in combination could distinguish a median lifespan up to 4.4 years shorter and a median lifespan at least 2.4 years shorter in an extensively adjusted model that controlled for various lifestyle factors, diagnosed medical conditions, and socioeconomic variables.

Furthermore, while neither the organizers of the HRS nor the authors returned any information to the respondents about their genetic predisposition, the polygenic scores were found to be associated with subjective life expectancy and self-rated health. This finding suggests that the unobserved genetic risks had actually been partly observed by the respondents, likely through their health or acquired medical conditions. Finally, greater genetic liability was found to be associated with several economic behaviors and measures, including 2.2 months shorter long-term care insurance coverage, but not with life insurance coverage.

### 1.1 Genetic testing, economic behavior, and insurance

The revelation of genetic risks may influence people’s expectations about their future health and longevity and, in turn, influence their economic behavior (Hamermesh 1985). The expected value of any kind of insurance tied to the health, life, or death of a person should theoretically be affected by knowledge of genetic risks, as should the rationale for purchasing coverage (Rothstein 2004; Peter, Richter, and Thistle 2017; Oster et al. 2010). If an applicant has private knowledge of risks that are not reflected in the premium (or benefit), then a particular insurance product will be considered either cheap or expensive (that is, actuarially unfair) depending on whether the benefit is paid specifically upon the death, survival, or illness of the insured. Thus, depending on the insurance product in question, it could be considered in the financial interest of an applicant to either withhold or reveal private knowledge of genetic risks when applying for insurance.

Only a handful of studies have empirically tested whether the decision to purchase insurance can be influenced by giving people novel information about a few of their genetic risk factors (Zick et al. 2000; Aktan-Collan, Haukkala, and Kääriäinen 2001; Armstrong et al. 2003; Zick et al. 2005; Taylor et al. 2010; Oster et al. 2010). While the results are mixed, some studies did indeed find that the willingness to purchase coverage increased among people who received a test result that indicated greater genetic risk, but this was true only for certain types of insurance. However, these studies were limited by their examination of self-reported rather than revealed preferences, their use of small sample sizes, their narrow focus on a few genetic variants or diseases, or the very limited insights into the role of genetics for common diseases that were available prior to 2010 (Visscher et al. 2017; Mills and Rahal 2019). Furthermore, it appears that the public’s genetic literacy is weak, even among the highly educated (Gericke et al. 2017; Chapman et al. 2019; Carver et al. 2017). Therefore, it is also conceivable that disregard for or misinterpretation of genetic test results leads to no behavioral changes at all or, alternatively, leads to changes that could be considered unexpected or inappropriate (Wang et al. 2018; Tandy-Connor et al. 2018; Nelson, Bowen, and Fullerton 2019; Lea et al. 2011). Thus, it remains an interesting and relevant empirical question to what extent health expectations and economic decisions, such as insurance purchases, are related to or influenced by the increasing availability of genetic information.

Representatives of various insurance providers have expressed concerns about the viability of different insurance products in a time of widespread genetic testing (Nabholz and Rechfeld 2017; Rechfeld et al. 2019; Hodgson and Haddow 2016). Their main worry is a situation in which insurance applicants with private knowledge of genetic risks would be sanctioned to withhold that information from underwriting (Rothstein 2004; Peter, Richter, and Thistle 2017). Theoretical analyses suggest that a ban on using genetic information for underwriting is Pareto suboptimal compared to alternative regulatory regimes of partial or full disclosure (Peter, Richter, and Thistle 2017). In addition, experts and stakeholders agree that sanctioned non-disclosure threatens the fundamental insurance principles of symmetric information and actuarial fairness, which can lead to adverse selection and escalating premiums (Harper 1993; Rothstein 2004; Nabholz and Somerville 2011). On the other hand, however, the danger of genetic discrimination could emerge if insurers were given access to genetic data, which could lead them to deny health care or financial support precisely to those individuals who need it most (Tiller et al. 2019; Newson et al. 2018). In some countries, there are reports of active genetic discrimination for some conditions (Joly, Ngueng Feze, and Simard 2013; Tiller et al. 2019).

Because of the real risk of genetic discrimination, many governments have taken a regulatory stance that favors consumer privacy over corporate interests by mostly limiting the rights of insurance providers to request and use genetic information to reject applications or determine premiums (Borry et al., 2012; Prince, 2019; Sijbrands et al., 2009). In countries that lack regulation, the insurance industry has frequently chosen to self-regulate with voluntary moratoriums. Over time, these developments could threaten the affordability and viability of several private insurance markets (Strohmenger and Wambach 2000; Hendren 2013), or in jurisdictions where insurance providers may request access to genetic data, providers could increase health or financial inequalities by discriminating on risk factors that are due to bad luck in the “genetic lottery” (Newson et al. 2018; Rothstein 2004). Therefore, the question of whether genetic test results should be disclosed to insurance providers is an urgent and controversial topic of societal relevance (Newson et al. 2018; Klitzman, Appelbaum, and Chung 2014). Recently, an international expert group of researchers and insurance stakeholders called for more research on this topic (Joly et al. 2014).

In practice, not all observable risks can be underwritten. Reasons include the negligible influence of certain risks on mortality or a lack of data to accurately define a fair premium (Nabholz and Somerville 2011). Thus, the accuracy and predictive scope of this new type of genetic information needs to be determined before polygenic scores can even be considered for use in underwriting. To date, only a handful of studies have investigated to what extent polygenic scores can classify people into groups of different mortality risks (Ganna et al. 2013; Marioni et al. 2016; Timmers et al. 2019; McDaid et al. 2017; Pilling et al. 2016; Joshi et al. 2017). The largest effect reported thus far is a 3.5-year difference in the median lifespan; this result was found by comparing the top versus bottom deciles of a polygenic score for parental lifespan. (Parental lifespan is a common proxy for individual longevity that imperfectly captures various diseases and lifestyle risks, among other factors.) However, estimates are still scarce, and most have been obtained without conditioning on observable confounders such as income, smoking, or medical conditions. Therefore, the main objective of this study is to estimate in another sample how well polygenic scores for common medical conditions and mortality risks can distinguish a difference in lifespan across groups of cumulative genetic liability and, in this regard, benchmark polygenic scores to conventional actuarial risk factors. In addition, as it is essentially unknown how much information polygenic scores can add on top of observable risk factors, we also investigate an extensively adjusted regression model.

Furthermore, we explore whether polygenic scores are associated with subjective life expectancy and self-rated health. Such an association may indicate whether the underlying genetic risks, which we assume are not directly observed by respondents, are nonetheless partly captured by these health measures. Finally, we investigate whether polygenic scores capture any differences in insurance purchases and other economically relevant variables in a situation where neither customers nor insurance providers have access to those data. We are aware of only a single recent study that tested for an association between polygenic scores for disease and economic behavior. Specifically, Shin, Lillard, and Bhattacharya (2019) tested whether a polygenic score for Alzheimer’s disease was associated with wealth composition in the HRS, and they found that greater genetic risk was associated with less wealth.

### 1.2 Genetic risks for common diseases

Genetic factors contribute substantially to the risk of disease (Visscher et al. 2017; Polderman et al. 2015). In a given population, genes may account for more than 30% of the variation in longevity (Pilling et al. 2017; Brooks-Wilson 2013; Ganna et al. 2013; Timmers et al. 2019). Diagnostic genetic tests for severe but rare single-gene disorders have been routine in clinical care for decades, for which thousands of tests are available (Phillips et al. 2018; Godard et al. 2003). However, most people are not affected by rare genetic disorders, and the contribution of these disorders to the mortality burden from noncommunicable diseases (NCDs) is limited, particularly in adults (Kaplan et al. 2013). Instead, a few common and substantially heritable medical conditions, such as cardiovascular disease (CVD), cancer, and diabetes, account for a majority of all NCD deaths (Khera et al. 2018; Jan et al. 2018; Bloom et al. 2011; Dagenais et al. 2019). A few prevalent, heritable, and potentially preventable mortality risks, such as high cholesterol and smoking, also cause a considerable NCD burden (Daar et al. 2007; Jan et al. 2018). Accordingly, the current study is restricted to medical conditions and mortality risks that are common in the population.

#### 1.2.1 Genome-wide association studies and polygenic scores

Common medical conditions are rarely, if ever, caused exclusively by any single gene. Instead, they are *complex traits* that are influenced by a large number of genetic variants with individually minute effects on disease risk (Dudbridge 2016; Visscher et al. 2017; Khera et al. 2018). This so-called *polygenicity* also applies to mortality risks and lifestyle factors (Willer et al. 2013; Karlsson Linnér et al. 2019; Dudbridge 2016). However, in combination, these small genetic effects sum up to the heritability of a trait, which accounts for ∼20–60% of the variance in longevity and many common diseases (Polderman et al. 2015; Witte, Visscher, and Wray 2014). In recent years, there has been rapid progress in the effort to identify genetic variants that contribute to complex traits. *Genome-wide association studies* (GWAS) are currently the method of choice for this purpose (Mills and Rahal 2019; Tam et al. 2019; Young et al. 2019).

A typical GWAS tests millions of single-nucleotide polymorphisms (SNPs), one at a time, for association with a trait (Pasaniuc and Price 2017). SNPs refer to a base pair at a particular location in a genome that varies among people, which is the most common form of genetic variation that exists. It is now relatively cheap and easy to measure millions of SNPs across a genome (e.g., using DNA extracted from saliva samples, which is then genotyped using high-throughput array technologies). Many recent GWAS have been performed in hundreds of thousands of participants, a few have been performed in more than a million participants, and larger studies are expected in the near future (Saunders et al. 2019; Mills and Rahal 2019; Tam et al. 2019). For example, the most recent GWAS on longevity studied more than five hundred thousand people (Timmers et al. 2019). To date, the GWAS literature has successfully linked tens of thousands of SNPs with hundreds of common diseases, health risks, and lifestyle behaviors, with a respectable replication record (Mills and Rahal 2019; Buniello et al. 2018; Tam et al. 2019; Young et al. 2019).

Estimated GWAS coefficients can be used to construct polygenic scores in hold-out samples that were not included in the GWAS (Pasaniuc and Price 2017). In essence, polygenic scores are simple linear combinations of a person’s genotype, weighed by each SNP’s trait-specific effect (Dudbridge 2013). These scores can have substantial predictive accuracy when the trait in question is heritable and the input SNP effects have been estimated in a well-powered GWAS (Torkamani and Topol 2019; Khera et al. 2018; Mavaddat et al. 2019). For example, recent studies show that polygenic scores can stratify a severalfold increased risk of disease, a capacity that is comparable to or better than that of clinical risk factors or monogenic mutations, such as those involved in familial hypercholesterolemia (Abraham et al. 2019; Khera et al. 2018; Escott-Price et al. 2017). The utility of the method is even greater when polygenic scores are analyzed jointly or together with other observable factors, such as family or medical history (Krapohl et al. 2017). In the future, polygenic scores may even substitute for expensive biomarkers or complement observable risk factors measured with imprecision (Torkamani, Wineinger, and Topol 2018).

### 2 Methods

### 2.1 Data

The analyses reported here were performed according to a preregistered analysis plan^a^. We analyzed the HRS, a longitudinal household survey of elderly Americans that has been conducted biannually since 1992 (Sonnega et al. 2014). The purpose of the HRS is to facilitate studies on how the socioeconomic environment is related to health and aging, for which study participants provide broad consent. Overall, there were 13 waves of data available, spanning the years 1992–2018. In 2006, the HRS also started collecting genotype data (Domingue et al. 2017). We analyzed the publicly available HRS Longitudinal File 2016 (v1), curated by the RAND Corporation, together with restricted-access genetic data that are available upon request from the National Center for Biotechnology Information (NCBI) database of Genotypes and Phenotypes (dbGaP) (Mailman et al. 2007).

The HRS Longitudinal File contains rich demographic data from the family; health and medical; education, occupation, income and wealth; and retirement domains. We extensively searched and narrowed down a selection of approximately 30 variables that we considered important to include as covariates. Because of the unbalanced panel structure and many missing observations, we did not implement a panel data model. Instead, to vastly increase the sample size, we collapsed the panel structure into a cross-section in the following way: binary, ordinal, and categorical variables were assigned the most frequently occurring value across the waves, and continuous variables were assigned the median. All dollar values were converted to 2016 prices. We report the full set of variables and sample descriptive statistics in **Table 1**.

**Table 1.**
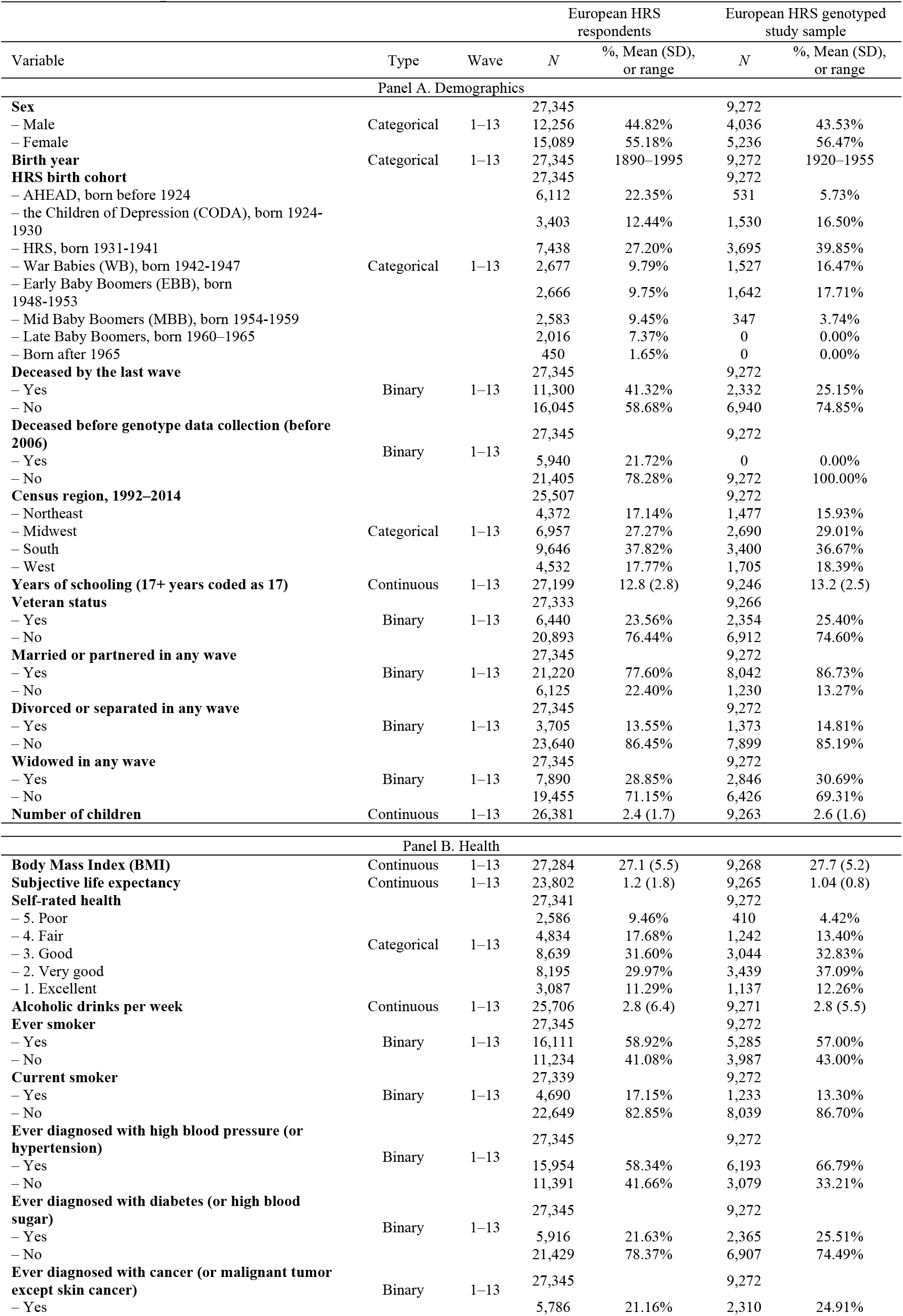
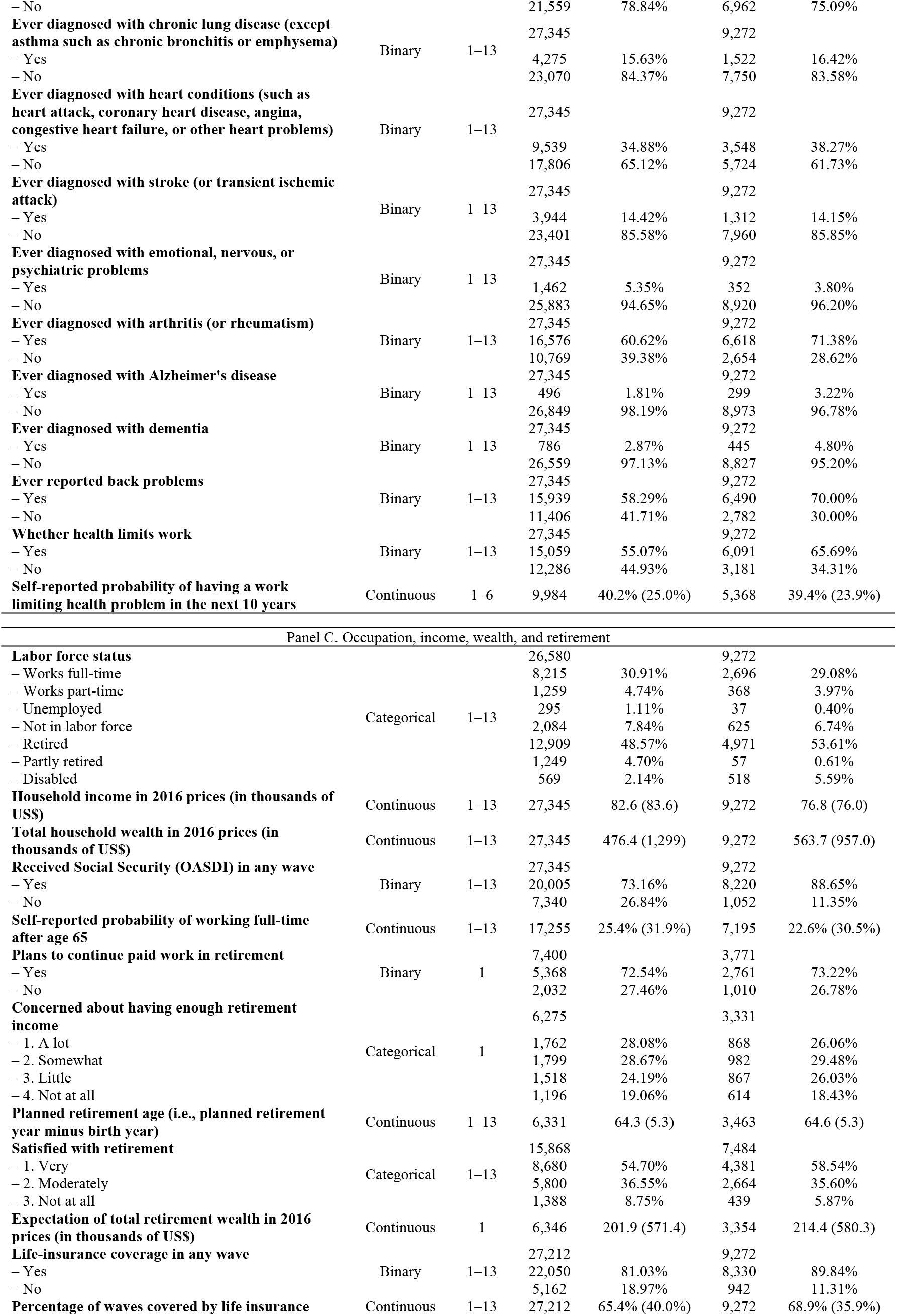
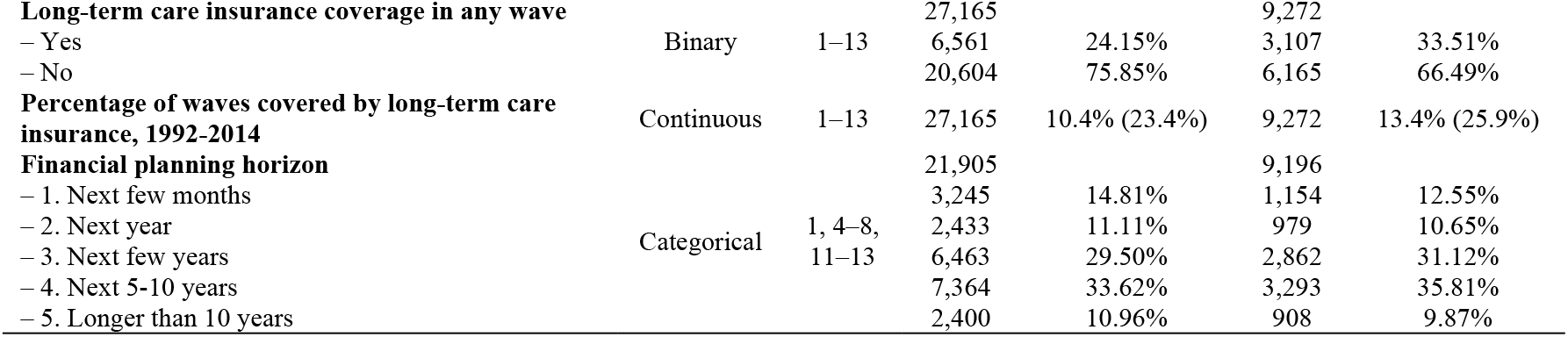
Descriptive statistics

#### 2.1.1 Respondent inclusion criteria

We restricted all analyses to respondents who self-reported that they categorize as both “White/Caucasian” and “Not Hispanic”, which we hereafter refer to as European ancestry^b^. The reason for this inclusion criterion is that the vast majority of GWAS have been performed in individuals of this descent, which drastically limits the possibility of constructing accurate polygenic scores in other ancestries (Duncan et al. 2019; Martin et al. 2017, 2019). In addition, in the HRS, this ancestral group is about three times larger than all the others combined. We analyzed the second release of the HRS genotype data, which had been imputed with the 1000 Genomes Project phase 1 version 3 reference panel (Auton et al. 2015). The second release includes 15,620 genotyped respondents, out of whom 10,958 reported that they were of European ancestry.

Next, we generated genetic principal components (PCs) to identify genetic outliers (Price et al. 2006, 2010). Specifically, we projected the European ancestry subsample of the 1000 Genomes phase 3 (version 5) reference panel onto the PCs and excluded respondents who had a value on any of the first four PCs that exceeded the range of the reference panel by more than 10%. In total, 10,701 individuals remained after this step. Thereafter, as is recommended (Price et al. 2006), we re-estimated 10 genetic PCs in the more homogenous HRS subsample, which were later included as covariates to control for population stratification. Population stratification refers to incidental covariation between allele frequencies and the outcome of interest, which can induce bias if not adjusted for (Price et al. 2006; Hamer and Sirota 2000). To be extra cautious, we excluded respondents who were more than 5 standard deviations from the mean on any of the newly generated PCs. This procedure removed 105 additional outliers. In total, 10,596 respondents remained at this stage. In summary, we proceeded conservatively to try to minimize the chance of bias, which can easily be introduced by ancestry admixture (Martin et al. 2017).

#### 2.1.2 Mortality selection and restriction of birth years

The HRS first started collecting genotype data in 2006, approximately 14 years after the first wave was collected. Thus, the genetic data could be subject to nonrandom selection based on mortality. In a thorough investigation of mortality selection in the HRS, Domingue et al. (2017) verified that the genotyped HRS respondents were indeed healthier, displayed fewer health-risk behaviors, and lived longer than the overall sample. We confirmed that there was mortality selection by comparing Kaplan-Meier survival functions (**Supplementary Figure 1**). Following recommendations given by Domingue et al., and with the aim to exclude birth years with fewer than a hundred observations with non-missing birth and death data, we restricted all further analyses to individuals born between 1920 and 1955. At this stage, we retained 9,272 respondents, of whom 2,332 were deceased by the most recent wave. We hereafter refer to these 9,272 genotyped respondents as our “study sample”. Any remaining mortality selection would most likely lead to an underestimation of the influence of polygenic scores, particularly for mortality risks that manifest before old age and that could have contributed to mortality selection, such as cardiovascular disease (Yusuf et al. 2019).

### Statistical analyses

#### 2.2.1 Quality control, heritability, and genetic correlations

We performed an extensive search of the published GWAS literature to identify the largest studies on common medical conditions and mortality risks that have thus far been made publicly available to the research community. The search was performed in the National Human Genome Research Institute (NGHRI) GWAS Catalog, which curates all published genome-wide association studies (MacArthur et al. 2017; Buniello et al. 2018), in the March 1, 2019, version of the database. To choose among the many hundreds of traits available in the database, we prespecified a selection that was guided by the medical literature. More specifically, we collected recognized predictors of mortality that had either been expert-curated by a panel of clinicians in a study by Ganna et al. (2013) or been determined to causally influence lifespan in a genetic study by McDaid et al. (2017). In addition, we prespecified the inclusion of only traits that had been studied in more than a hundred thousand people, since the accuracy of polygenic scores is a strong function of GWAS sample size (Daetwyler, Villanueva, and Woolliams 2008).

As reported in **Table 2**, we collected results from 29 GWAS (two were excluded see below) that spanned many medical domains, including cardiology, oncology, neurology, and psychiatry; in total 15 common medical conditions and the following 14 mortality risks: three measures of blood pressure, body mass index (BMI), four measures of blood cholesterol, educational attainment, height, parental lifespan, smoking initiation and cigarettes per day (smoking intensity), and (alcoholic) drinks per week. The average sample size was ∼455,000, and the largest was above a million (atrial fibrillation).

**Table 2.**
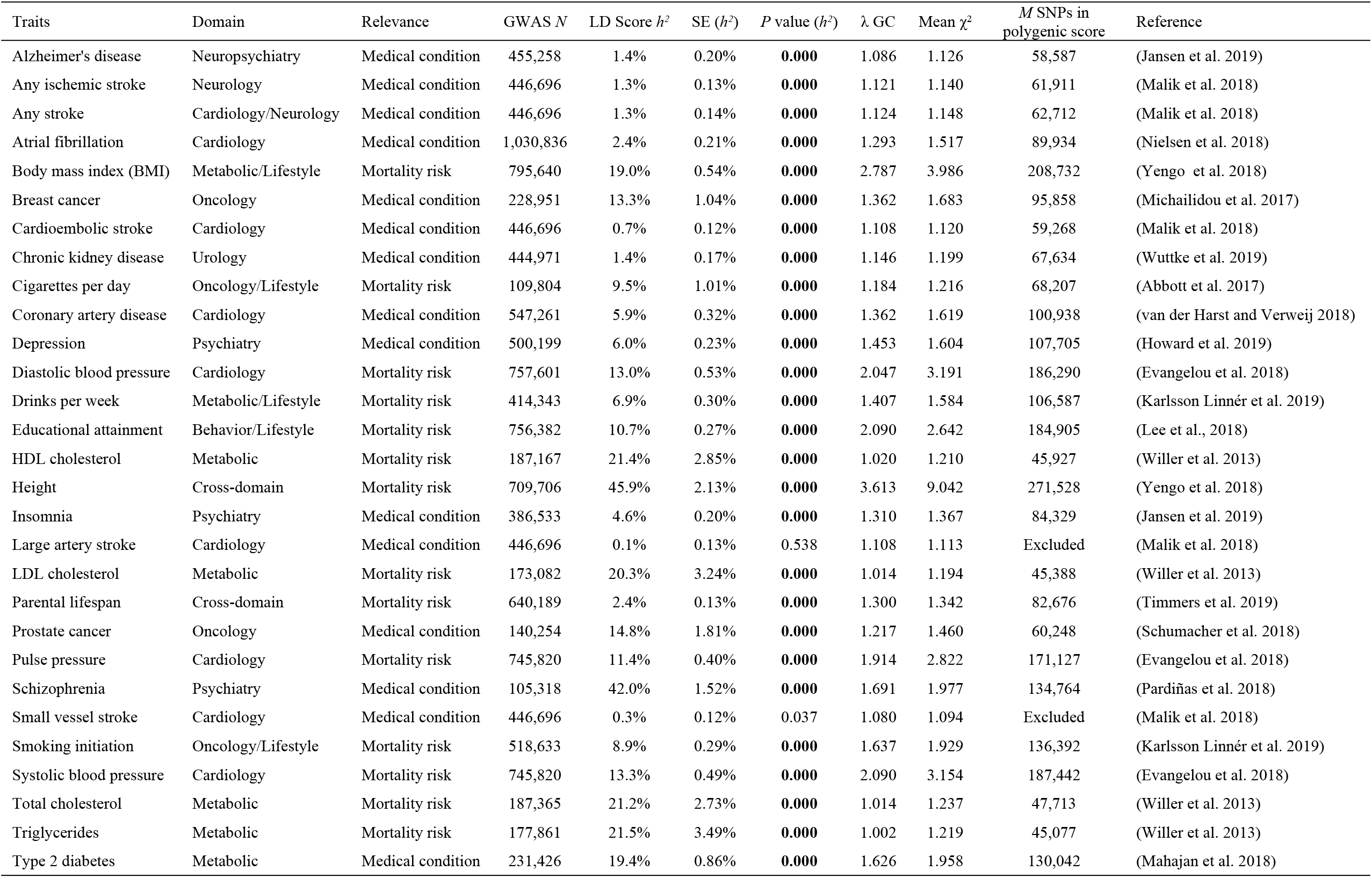
GWAS results for common medical conditions and mortality risks

| Traits | Domain | Relevance | GWAS $N$ | LD Score $h^2$ | SE ($h^2$) | $P$ value ($h^2$) | $\lambda$ GC | Mean $\chi^2$ | $M$ SNPs in polygenic score | Reference |
| --- | --- | --- | --- | --- | --- | --- | --- | --- | --- | --- |
| Alzheimer's disease | Neuropsychiatry | Medical condition | 455,258 | 1.4% | 0.20% | **0.000** | 1.086 | 1.126 | 58,587 | (Jansen et al. 2019) |
| Any ischemic stroke | Neurology | Medical condition | 446,696 | 1.3% | 0.13% | **0.000** | 1.121 | 1.140 | 61,911 | (Malik et al. 2018) |
| Any stroke | Cardiology/Neurology | Medical condition | 446,696 | 1.3% | 0.14% | **0.000** | 1.124 | 1.148 | 62,712 | (Malik et al. 2018) |
| Atrial fibrillation | Cardiology | Medical condition | 1,030,836 | 2.4% | 0.21% | **0.000** | 1.293 | 1.517 | 89,934 | (Nielsen et al. 2018) |
| Body mass index (BMI) | Metabolic/Lifestyle | Mortality risk | 795,640 | 19.0% | 0.54% | **0.000** | 2.787 | 3.986 | 208,732 | (Yengo et al. 2018) |
| Breast cancer | Oncology | Medical condition | 228,951 | 13.3% | 1.04% | **0.000** | 1.362 | 1.683 | 95,858 | (Michailidou et al. 2017) |
| Cardioembolic stroke | Cardiology | Medical condition | 446,696 | 0.7% | 0.12% | **0.000** | 1.108 | 1.120 | 59,268 | (Malik et al. 2018) |
| Chronic kidney disease | Urology | Medical condition | 444,971 | 1.4% | 0.17% | **0.000** | 1.146 | 1.199 | 67,634 | (Wuttke et al. 2019) |
| Cigarettes per day | Oncology/Lifestyle | Mortality risk | 109,804 | 9.5% | 1.01% | **0.000** | 1.184 | 1.216 | 68,207 | (Abbott et al. 2017) |
| Coronary artery disease | Cardiology | Medical condition | 547,261 | 5.9% | 0.32% | **0.000** | 1.362 | 1.619 | 100,938 | (van der Harst and Verweij 2018) |
| Depression | Psychiatry | Medical condition | 500,199 | 6.0% | 0.23% | **0.000** | 1.453 | 1.604 | 107,705 | (Howard et al. 2019) |
| Diastolic blood pressure | Cardiology | Mortality risk | 757,601 | 13.0% | 0.53% | **0.000** | 2.047 | 3.191 | 186,290 | (Evangelou et al. 2018) |
| Drinks per week | Metabolic/Lifestyle | Mortality risk | 414,343 | 6.9% | 0.30% | **0.000** | 1.407 | 1.584 | 106,587 | (Karlsson Linnér et al. 2019) |
| Educational attainment | Behavior/Lifestyle | Mortality risk | 756,382 | 10.7% | 0.27% | **0.000** | 2.090 | 2.642 | 184,905 | (Lee et al., 2018) |
| HDL cholesterol | Metabolic | Mortality risk | 187,167 | 21.4% | 2.85% | **0.000** | 1.020 | 1.210 | 45,927 | (Willer et al. 2013) |
| Height | Cross-domain | Mortality risk | 709,706 | 45.9% | 2.13% | **0.000** | 3.613 | 9.042 | 271,528 | (Yengo et al. 2018) |
| Insomnia | Psychiatry | Medical condition | 386,533 | 4.6% | 0.20% | **0.000** | 1.310 | 1.367 | 84,329 | (Jansen et al. 2019) |
| Large artery stroke | Cardiology | Medical condition | 446,696 | 0.1% | 0.13% | 0.538 | 1.108 | 1.113 | Excluded | (Malik et al. 2018) |
| LDL cholesterol | Metabolic | Mortality risk | 173,082 | 20.3% | 3.24% | **0.000** | 1.014 | 1.194 | 45,388 | (Willer et al. 2013) |
| Parental lifespan | Cross-domain | Mortality risk | 640,189 | 2.4% | 0.13% | **0.000** | 1.300 | 1.342 | 82,676 | (Timmers et al. 2019) |
| Prostate cancer | Oncology | Medical condition | 140,254 | 14.8% | 1.81% | **0.000** | 1.217 | 1.460 | 60,248 | (Schumacher et al. 2018) |
| Pulse pressure | Cardiology | Mortality risk | 745,820 | 11.4% | 0.40% | **0.000** | 1.914 | 2.822 | 171,127 | (Evangelou et al. 2018) |
| Schizophrenia | Psychiatry | Medical condition | 105,318 | 42.0% | 1.52% | **0.000** | 1.691 | 1.977 | 134,764 | (Pardiñas et al. 2018) |
| Small vessel stroke | Cardiology | Medical condition | 446,696 | 0.3% | 0.12% | 0.037 | 1.080 | 1.094 | Excluded | (Malik et al. 2018) |
| Smoking initiation | Oncology/Lifestyle | Mortality risk | 518,633 | 8.9% | 0.29% | **0.000** | 1.637 | 1.929 | 136,392 | (Karlsson Linnér et al. 2019) |
| Systolic blood pressure | Cardiology | Mortality risk | 745,820 | 13.3% | 0.49% | **0.000** | 2.090 | 3.154 | 187,442 | (Evangelou et al. 2018) |
| Total cholesterol | Metabolic | Mortality risk | 187,365 | 21.2% | 2.73% | **0.000** | 1.014 | 1.237 | 47,713 | (Willer et al. 2013) |
| Triglycerides | Metabolic | Mortality risk | 177,861 | 21.5% | 3.49% | **0.000** | 1.002 | 1.219 | 45,077 | (Willer et al. 2013) |
| Type 2 diabetes | Metabolic | Medical condition | 231,426 | 19.4% | 0.86% | **0.000** | 1.626 | 1.958 | 130,042 | (Mahajan et al. 2018) |

Next, we performed quality control to exclude rare and low-quality SNPs (minor allele frequency less than 0.01 and imputation quality less than 0.9), in accordance with the standards of the field (Winkler et al. 2014). It is recommended to exclude such SNPs to increase the signal-to-noise ratio in polygenic scores (Kuchenbaecker et al. 2017). We estimated SNP heritabilities (the proportion of variation explained by a set of SNPs) by applying LD Score regression on the GWAS results (Bulik-Sullivan, Loh, et al. 2015; Bulik-Sullivan, Finucane, et al. 2015). The method utilizes the fact that under a polygenic model, genetic variants that are correlated—in *linkage disequilibrium* (LD)—with many other variants are more likely to capture genetic signals across a genome. A variant’s LD Score, the sum of its squared correlations with other variants, has a proportional relationship with the expectation of its association test statistic. This relationship can be transformed to an approximate lower-bound SNP heritability and to test for population stratification under various conditions (Lee, McGue, et al. 2018).

To eliminate GWAS with negligible genetic signals, we excluded two traits for which LD Score heritability was not distinguishable from zero at the preregistered threshold *P* value less than 0.001: (1) large artery stroke (*h*^*2*^ = 0.07%; *P* = 0.59) and (2) small vessel stroke (*h*^*2*^ = 0.25%; *P* = 0.037). The heritability of the 27 remaining traits ranged from 0.7% (cardioembolic stroke) to 45.9% (height). Next, we estimated pairwise LD Score genetic correlations (*r*_*g*_) (Bulik-Sullivan, Finucane, et al. 2015) (**Supplementary Table 1** and **Supplementary Figure 2**). The method estimates genetic correlations by utilizing the overlap in association test statistics across SNPs as a measure of genetic covariance while adjusting for LD and sample overlap. It has been shown that the method can robustly estimate genetic overlap across a range of plausible confounding scenarios (Lee, McGue, et al. 2018). Importantly, we found that most of the common medical conditions and mortality risks we collected were moderately genetically correlated with parental lifespan, which suggests that they should be able to capture variation in survival (Timmers et al. 2019; McDaid et al. 2017; Marioni et al. 2016; Daetwyler, Villanueva, and Woolliams 2008).

#### 2.2.2 Polygenic scores

In our study sample, we computed polygenic scores as linear combinations of individual-level genotypes weighed by trait-specific GWAS effects. Thus, polygenic scores aggregate an individual’s genetic liability (or propensity) towards a trait or disease into a genetic predictor (Dudbridge 2013; Khera et al. 2018; Torkamani, Wineinger, and Topol 2018). The *i*th respondent’s polygenic score for the *k*th trait, ***S***_*ik*_, was computed as

**Table 3.** Selection of hierarchical Cox regression results^1^

|  | Model 3 (*N* = 9,246; deaths = 2,321) | Model 4 (*N* = 9,007; deaths = 2,223) |
| --- | --- | --- |
| Regressors (polygenic scores shown in bold) | Coefficient (Robust SE) | Coefficient (Robust SE) |
| **Alzheimer's disease** | 0.052 (0.023)* | 0.057 (0.024)* |
| **Atrial fibrillation** | 0.054 (0.023)* | 0.046 (0.025) † |
| **Cigarettes per day (smoking intensity)** | 0.073 (0.023)*** | 0.064 (0.024)** |
| **Height** | 0.049 (0.025)* | 0.045 (0.026) |
| **Type 2 diabetes** | 0.054 (0.026)* | 0.029 (0.028) |
| **Parental lifespan** | -0.087 (0.025)*** | -0.058 (0.028)* |
| Sex (0 = Male; 1 = Female) | -0.872 (0.241)*** | -0.568 (0.302) † |
| Census region (base: Northeast) |  |  |
| – Midwest | -0.107 (0.072) | -0.035 (0.075) |
| – South | 0.048 (0.068) | 0.034 (0.072) |
| – West | -0.063 (0.080) | 0.023 (0.083) |
| Years of schooling | -0.058 (0.010)*** | -0.024 (0.010)* |
| Log of total household income (2016 prices) | -0.201 (0.046)*** | -0.089 (0.047) † |
| Labor force participation (base: Works full-time) |  |  |
| – Works part-time | -0.167 (0.169) | -0.066 (0.165) |
| – Unemployed | 0.276 (0.532) | 0.243 (0.552) |
| – Partly retired | -0.335 (0.111)** | -0.355 (0.118)** |
| – Retired | -0.251 (0.073)*** | -0.385 (0.076)*** |
| – Disabled | 1.119 (0.270)*** | 0.725 (0.314)* |
| – Not in labor force | -0.148 (0.117) | -0.198 (0.118) † |
| Received “Social Security” (OASDI) in any wave | -0.430 (0.117)*** | -0.496 (0.124)*** |
| Married or partnered in any wave | -0.200 (0.071)** | -0.158 (0.072)* |
| Widowed in any wave | -0.357 (0.056)*** | -0.348 (0.057)*** |
| Sex-specific birth-year dummies | YES | YES |
| Birth-month dummies | YES | YES |
| Subjective life expectancy |  | -0.086 (0.029)** |
| Self-rated health (Base: 1. Excellent) |  |  |
| – 2. Very good |  | -0.050 (0.089) |
| – 3. Good |  | 0.304 (0.091)*** |
| – 4. Fair |  | 0.633 (0.102)*** |
| – 5. Poor |  | 1.069 (0.134)*** |
| BMI |  | 0.013 (0.006)* |
| Alcoholic drinks per week |  | 0.012 (0.005)* |
| Current smoker |  | 0.812 (0.071)*** |
| Ever smoker |  | 0.193 (0.053)*** |
| Maternal max attained age |  | -0.003 (0.002) † |
| Paternal max attained age |  | -0.003 (0.002)* |
| Ever diagnosed with high blood pressure |  | -0.022 (0.054) |
| Ever diagnosed with diabetes |  | 0.136 (0.054)* |
| Ever diagnosed with cancer |  | 0.244 (0.049)*** |
| Ever diagnosed with chronic lung disease |  | 0.348 (0.058)*** |
| Ever diagnosed with heart conditions |  | 0.069 (0.049) |
| Ever diagnosed with stroke |  | 0.091 (0.058) |
| Ever diagnosed with psychiatric disorder |  | 0.082 (0.105) |
| Ever diagnosed with arthritis |  | -0.165 (0.056)** |
| Ever diagnosed with Alzheimer's disease |  | 0.208 (0.090)* |
| Ever diagnosed with dementia |  | -0.180 (0.082)* |
| Ever diagnosed with back problems |  | -0.356 (0.053)*** |
| Global test if model violates PH assumption | $\chi^2(128\text{ df}) = 131.0; P = 0.404$ | $\chi^2(150\text{ df}) = 203.0; P < 0.001$ |
| Cox-Snell *R*<sup>2</sup> | 0.231 | 0.286 |
| Harrel's *c* statistic | 0.816 (0.004) | 0.850 (0.004) |
| Gönen & Heller's *K* statistic | 0.857 (0.003) | 0.862 (0.003) |

Notes: † $P \leq 0.1$ ; \* $P \leq 0.05$ ; \*\* $P \leq 0.01$ ; \*\*\* $P \leq 0.001$ . Robust standard errors were clustered at the household level.
<sup>1</sup> The complete results are reported in **Supplementary Table 3**.

**Table 4.**
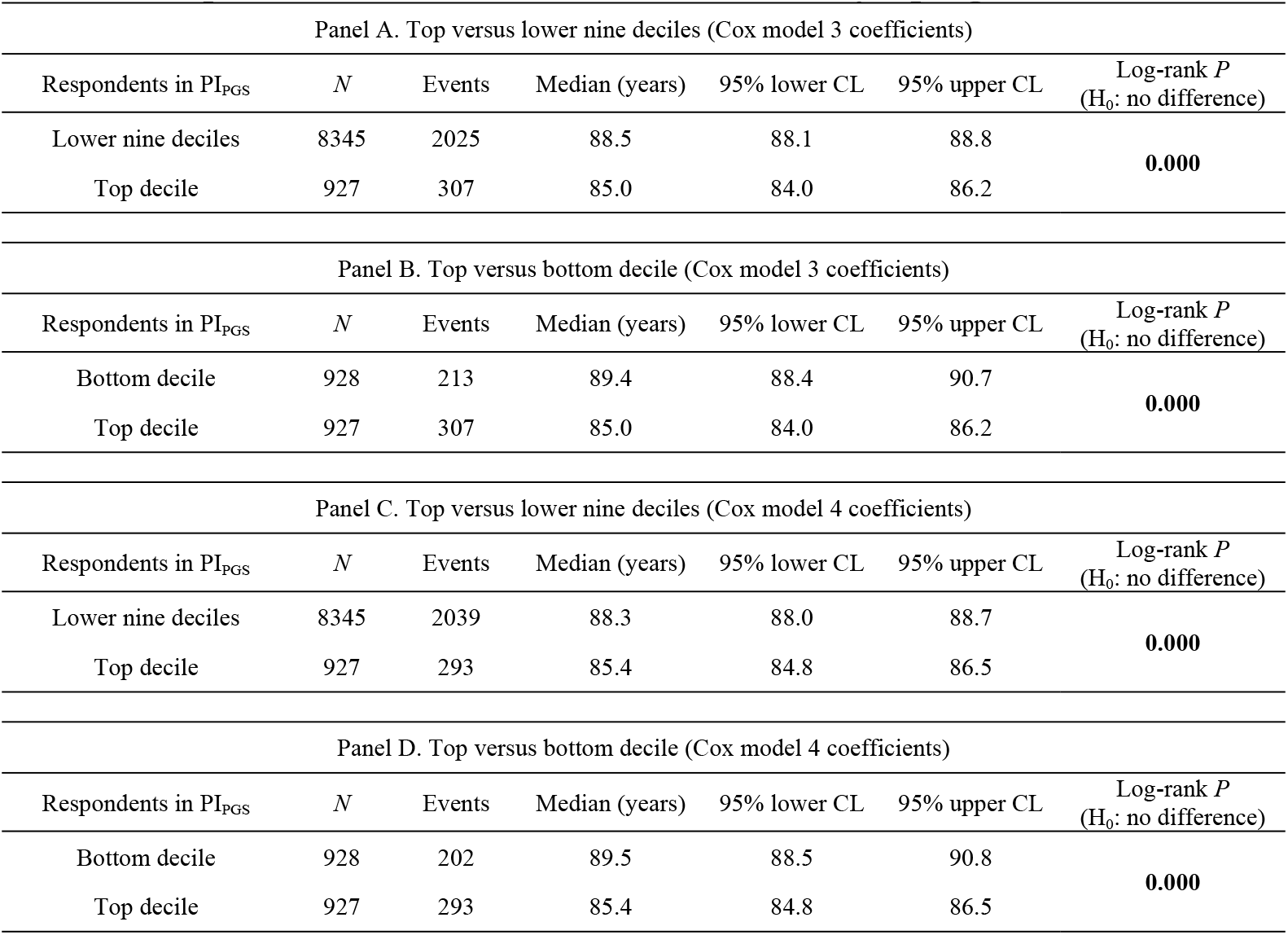
Kaplan-Meier survival estimates stratified by a prognostic index (PI)

**Table 5.** Kaplan-Meier survival estimates stratified by the prognostic indices (PIs)

| Panel A. Top versus lower nine deciles of the PIs (computed with model 3 coefficients) |  |  |  |  |  |  |  |  |
| --- | --- | --- | --- | --- | --- | --- | --- | --- |
| Respondents in<br>PI<sub>PGS</sub> | Respondents in<br>PI<sub>PC</sub> | Respondents in<br>PI<sub>COVAR</sub> | *N* | Events | Median<br>(years) | 95%<br>lower<br>CL | 95%<br>upper<br>CL | Log-rank *P*<br>(H<sub>0</sub>: no<br>difference) |
| Lower nine deciles | Lower nine deciles | Lower nine deciles | 6764 | 1571 | 88.9 | 88.6 | 89.3 | **0.000** |
| Top decile | Lower nine deciles | Lower nine deciles | 736 | 236 | 85.8 | 84.6 | 86.7 |  |
| Top decile | Top decile | Lower nine deciles | 82 | 29 | 84.2 | 81.2 | 89.5 |  |
| Top decile | Top decile | Top decile | 10 | 3 | 80.8 | 59.4 | -- |  |
| Lower nine deciles | Top decile | Lower nine deciles | 740 | 204 | 88.0 | 86.7 | 88.6 |  |
| Lower nine deciles | Top decile | Top decile | 94 | 28 | 79.0 | 77.7 | 85.2 |  |
| Lower nine deciles | Lower nine deciles | Top decile | 723 | 211 | 85.0 | 83.2 | 86.2 |  |
| Top decile | Lower nine deciles | Top decile | 97 | 39 | 80.8 | 74.0 | 82.4 |  |
| Panel B. Top versus lower nine deciles of the PIs (computed with model 4 coefficients) |  |  |  |  |  |  |  |  |
| Respondents in<br>PI<sub>PGS</sub> | Respondents in<br>PI<sub>PC</sub> | Respondents in<br>PI<sub>COVAR</sub> | *N* | Events | Median<br>(years) | 95%<br>lower<br>CL | 95%<br>upper<br>CL | Log-rank *P*<br>(H<sub>0</sub>: no<br>difference) |
| Lower nine deciles | Lower nine deciles | Lower nine deciles | 6605 | 1419 | 89.2 | 88.8 | 89.6 | **0.000** |
| Top decile | Lower nine deciles | Lower nine deciles | 694 | 195 | 86.8 | 85.4 | 87.8 |  |
| Top decile | Top decile | Lower nine deciles | 86 | 28 | 85.9 | 83.9 | 89.5 |  |
| Top decile | Top decile | Top decile | 11 | 6 | 70.7 | 63.6 | -- |  |
| Lower nine deciles | Top decile | Lower nine deciles | 722 | 170 | 88.8 | 87.6 | 89.6 |  |
| Lower nine deciles | Top decile | Top decile | 83 | 45 | 76.3 | 75.8 | 78.7 |  |
| Lower nine deciles | Lower nine deciles | Top decile | 696 | 309 | 78.5 | 77.3 | 79.8 |  |
| Top decile | Lower nine deciles | Top decile | 110 | 51 | 74.4 | 72.7 | 76.4 |  |

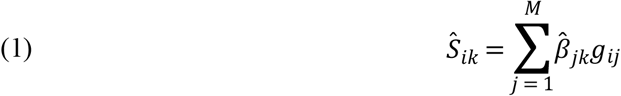

where the respondent’s genotype *g*_*ij*_ at SNP *j* was weighed by the corresponding trait-specific GWAS effect, 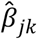, and then summed across *M* SNPs. In the regression analyses below, we entered multiple polygenic scores on the right-hand side of the regression equation, which can increase the predictive accuracy of genetically correlated outcomes (Krapohl et al. 2017).

We excluded weak GWAS associations at a *P* value greater than 0.05 to reduce noise from estimation error (Choi and O’Reilly 2019) and then constructed polygenic scores using the remaining SNPs that overlapped with the 1.4 million SNPs in the high-quality consensus genotype set established by the International HapMap 3 Consortium (Altshuler et al. 2010; Chang et al. 2015). This is a common approach that achieves reasonable genome-wide coverage with high imputation accuracy without including too many correlated SNPs, which can reduce predictive accuracy (Karlsson Linnér et al. 2019; Lee, Wedow, et al. 2018). The final number of SNPs in the scores ranged from 45,077 to 271,528 (**Table 2**). HRS participants did not obtain any information about their polygenic values.

#### 2.2.3 Univariate survival analysis and Cox proportional hazards regression

We first performed a series of nonparametric univariate analyses of respondent, maternal, and paternal survival by estimating stratified Kaplan-Meier survival functions (**Supplementary Table 2**) (Kaplan and Meier 1958; Clark et al. 2003). A benefit of analyzing parental survival is reduced censoring and mortality selection, but a disadvantage is that offspring genotypes are a noisy measure of parental genotypes (Wright et al. 2019). Additionally, mortality risks change over time, and it is questionable whether the same genetic and environmental risks remain relevant. We analyzed monthly survival in the respondents but observed only yearly survival in the parents.

While the univariate analysis can suggest factors that appear to be associated with survival, it is not statistically conditioned on other variables (Hosmer and Lemeshow 1998). Therefore, we proceeded by performing multiple regression of respondent survival by estimating a series of Cox proportional hazards (PH) models (**Table 3** and **Supplementary Table 3**) (Bradburn et al. 2003a; Royston and Altman 2013). We did not perform Cox regression of parental survival because we could not observe its covariates. In these analyses, we included 10 standardized genetic PCs. Because of the HRS household structure, we clustered the standard errors at the household level. We estimated four nested regression models hierarchically. In summary, the models included the following regressors in addition to the genetic PCs:

1. all polygenic scores except the score for parental lifespan;
2. model (1) together with sex-specific birth-year dummies, birth-month dummies, and demographic and socioeconomic covariates, including years of schooling and income;
3. model (2) together with the polygenic score for parental lifespan; and
4. model (3) together with covariates from the health and lifestyle risk domains, including BMI, smoking, drinking, parental lifespan, subjective life expectancy, self-rated health, and 11 indicators for categories of diagnosed medical conditions.

Overall, we consider model (3) to be our preferred model for the development of a prognostic index of genetic risk that could be evaluated early in life before any signs or symptoms of disease emerge (see below). At the same time, model (4) indicates whether polygenic scores have any capacity to distinguish lifespan above and beyond the inclusion of intermediate variables that lie on the causal pathway between genetic risk and mortality, such as manifested medical conditions.

#### 2.2.4 Model diagnostics and fit

We evaluated a series of model diagnostics for the Cox models (Moore 2016; Bradburn et al. 2003b; Klein and Moeschberger 2003). First, we checked whether the models or any of the regressors violated the PH assumption by testing the scaled Schoenfeld residuals for association with time to event. Second, we examined whether we had chosen a suitable functional form for the covariates by visually inspecting their relationship with the Martingale residuals. Third, we examined the deviance residuals for outliers or influential observations.

We then assessed the model fit. We computed likelihood-ratio tests, Wald tests, and log-rank tests to evaluate whether the regressors improved the model fit above the null model. Next, we computed the Cox-Snell pseudo-*R*^*2*^, Harrel’s *c* statistic, and Gönen & Heller’s *K* statistic (Harrell Jr et al. 1982). The latter two are concordance measures that compare the observed time to event with the ranks of the respondents’ hazards as predicted by the fitted model, akin to an area under the ROC curve (AUC) measure (Royston and Altman 2013). In the next section, we explain how we computed the Royston & Sauerbrei 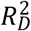 measure of model fit. Finally, we performed likelihood-ratio tests between the nested models to examine whether model fit improved. Because of the large number of events, *N*_deceased_ = 2,332, we performed no stepwise covariate selection.

#### 2.2.5 Prognostic indices of survival

To investigate how well the polygenic scores, when combined, could stratify survival relative to (i) genetic PCs and (ii) covariates, we computed three prognostic indices (PI) for each of the four Cox models. In this context, a PI is a weighted sum of multiple variables, which are weighed by their Cox coefficients. In other words, we aggregated the influence of sets of variables into hazard indices. The *i*th respondent’s PI across the polygenic scores was computed as

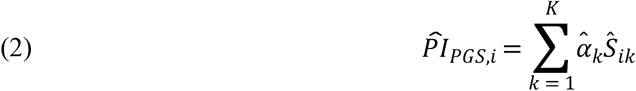

where the respondent’s polygenic score, *Ŝ*_*ik*_, was weighed by its Cox regression coefficient, 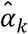, and then summed across the *K* polygenic scores. The PIs for the genetic PCs (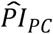) and the covariates (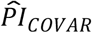) were computed analogously, with the exception that 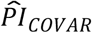 excluded the sex-specific birth-year dummies and the birth-month dummies, because we considered those to capture time and sampling effects rather than meaningful individual differences.

To evaluate the relative variance explained by the three 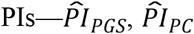, and 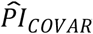—we computed the Royston & Sauerbrei 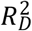 measure of model fit (Royston and Sauerbrei 2004; Royston and Altman 2013; Bradburn et al. 2003b). This method orders respondents according to a PI and then projects them onto a normal distribution to attain so-called rankits (expected *Z*-scores based on the order and number of the individuals). Then, an auxiliary Cox regression is performed on the rankits alone, and the resulting regression coefficient can be transformed into a measure of explained variation on the log hazard scale. Thus, 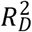 is more similar to the traditional coefficient of determination (*R*^*2*^) of linear regression than to the Cox-Snell pseudo-*R*^*2*^, which is computed as ratios of log-likelihoods. We performed this comparison across all four Cox models (**Supplementary Table 3**).

#### 2.2.6 Comparison of stratified survival functions

Next, for Cox models (3) and (4), we compared stratified survival functions in two ways: the first comparison (a) was stratified by the top versus lower nine deciles of the PI distribution, and the second comparison (b) was stratified by the top versus bottom deciles. Comparison (a) was deliberately chosen to mirror medical underwriting, where primarily individuals with a substantially increased risk are classified as substandard and charged a higher premium (Nabholz and Somerville 2011; Rothstein 2004), while comparison (b) was similar to a traditional extreme-groups approach (Preacher et al. 2005; Timmers et al. 2019). We report both comparisons but focus our discussion on the first comparison (a), which did not discard any data. In these analyses, the 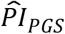 based on model (3) was our preferred genetic predictor of interest (**Table 4** and **Figure 1**). We performed a log-rank test to determine whether the survival functions of the two groups in each comparison differed, and we evaluated the size of the difference in median lifespan (Clark et al. 2003). Thereafter, we stratified survival by the three PIs simultaneously but only for comparison (a), as simultaneously stratifying by the top and bottom deciles of the three PIs would include too few respondents (**Table 5** and **Figure 2**).

**Figure 1.**
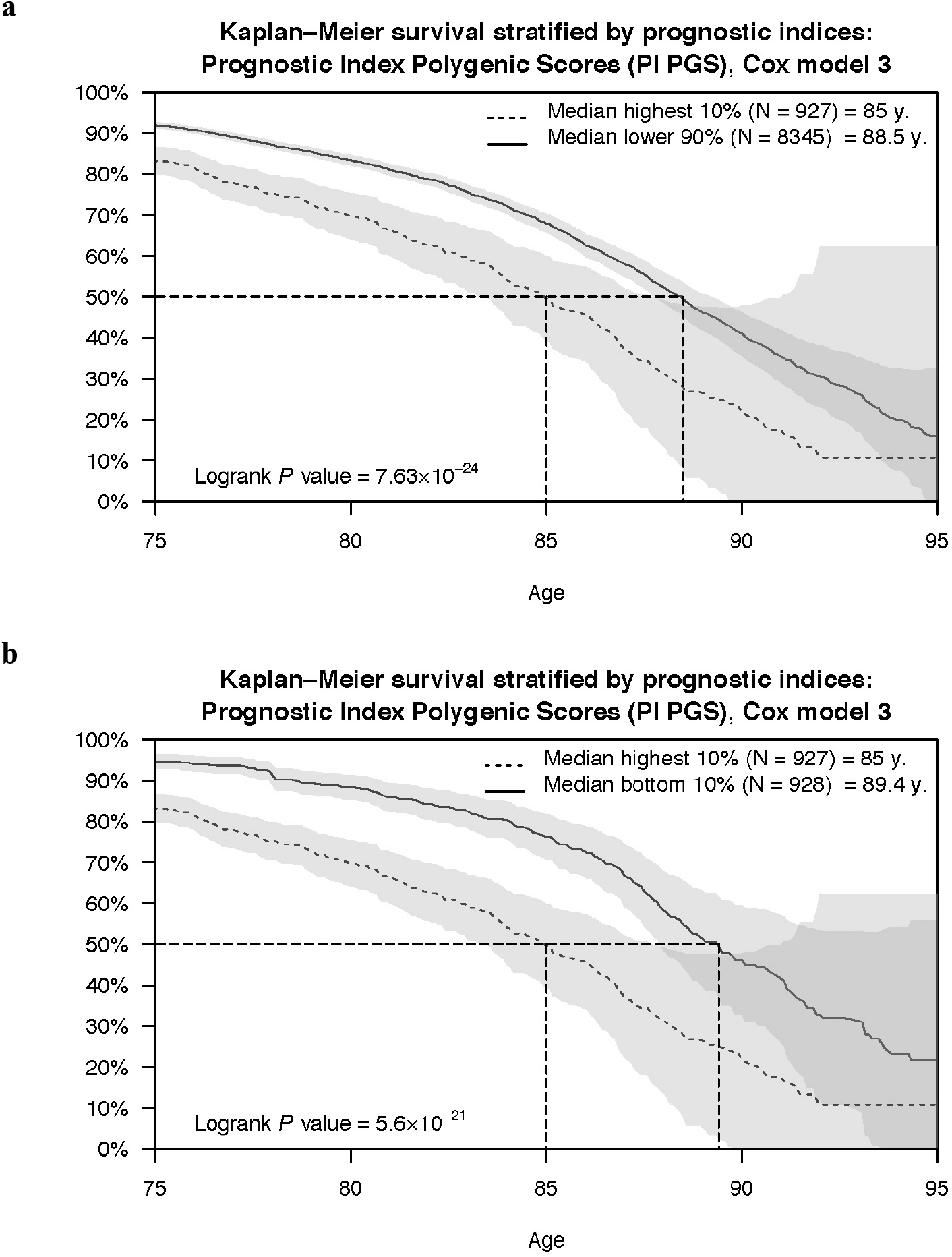
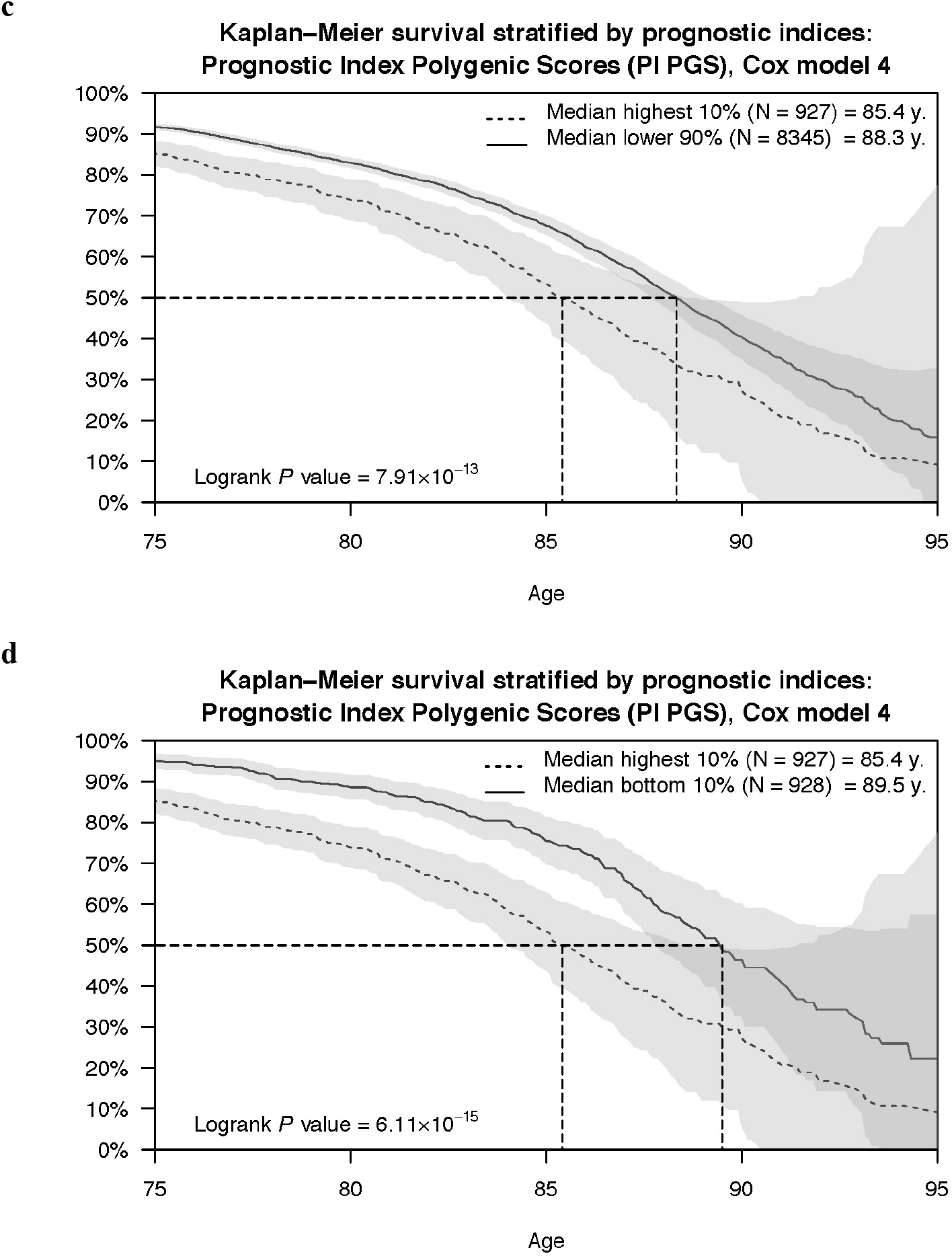
Kaplan-Meier survival curves stratified by the prognostic index of the polygenic scores. Using the prognostic index of the polygenic scores (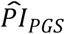) computed with the coefficient estimates of Cox models (3) and (4), we performed two comparisons of stratified survival functions. The first comparison (panels **a** and **c**) was stratified by the top versus lower nine deciles of the PI distribution, while the second comparison (panels **b** and **d**) was stratified by the top versus bottom decile. The dashed lines display the median survival in the two strata. The log-rank *P* value indicates whether the survival functions (not the median) of the two strata are different.

**Figure 2.**
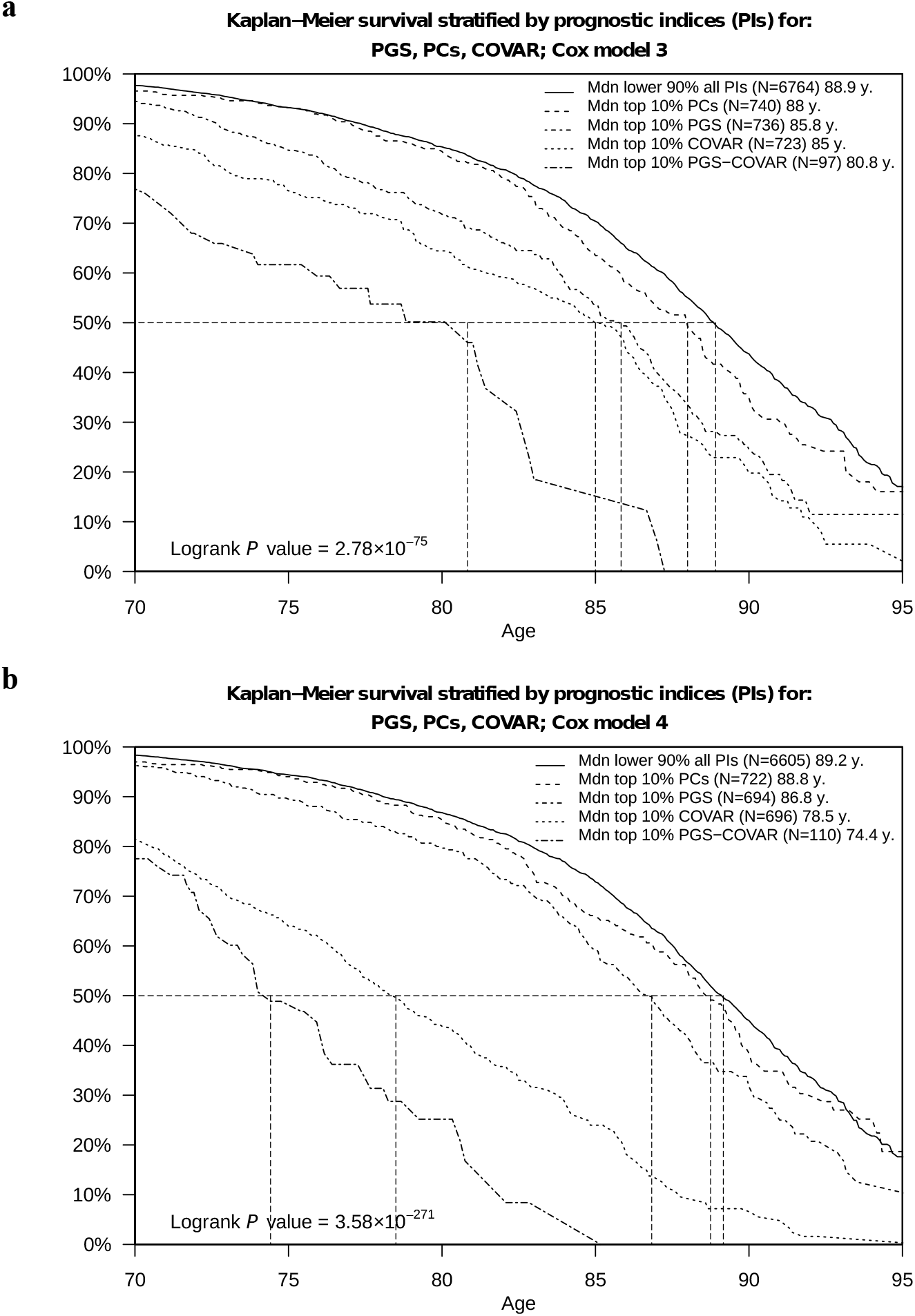
Kaplan-Meier survival curves stratified by the three prognostic indices. Using the prognostic indices of the polygenic scores (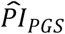), the genetic PCs (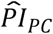), and the covariates (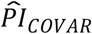), computed with the coefficient estimates of Cox models 3 (panel **a**) and 4 (panel **b**), we compared the simultaneously stratified survival functions by the top versus lower nine deciles of the distribution of the three PIs. The dashed lines display the median survival in the strata. The log-rank *P* value indicates whether the survival functions (not the median) of the strata are different. Three strata are not displayed in the figure but instead in **Table 5**.

#### 2.2.7 Benchmark to conventional actuarial risk factors

To benchmark the 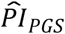, we stratified Kaplan-Meier survival functions by each of the following conventional actuarial risk factors (**Supplementary Table 4** and **Supplementary Figure 6**): smoking (never, current, and former); BMI; years of schooling; log of household income; sex; and ever diagnosed with (a) high blood pressure (or hypertension), (b) diabetes (or high blood sugar), (c) cancer (or malignant tumor except skin cancer), (d) chronic lung disease (except asthma such as chronic bronchitis or emphysema), (e) heart conditions (such as heart attack or coronary heart disease), or (f) stroke (or transient ischemic attack). To avoid mortality selection from the genotyping procedure, we performed this analysis in both our study sample and the full sample of 27,345 European HRS respondents.

#### 2.2.8 Cross-validation of the preferred Cox model

To maximize the sample size, we analyzed the PIs in the same sample as the one used to estimate the Cox coefficients. Therefore, we performed a cross-validation with 1,000 iterations for our preferred model (3) to evaluate the possibility of overfitting (Royston and Sauerbrei 2004). In each iteration, we trained the coefficients of the 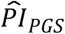 in a random sample containing 65% of the households (*H*_training_ = 4,325) and used the remaining households as a validation sample (*H*_validation_ = 2,329). First, we evaluated the so-called “calibration slope” by fitting the 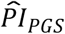 in model (3) instead of the polygenic scores (Royston and Altman 2013). If the slope is different from one, then the cross-validation discrimination is either better (>1) or worse (<1). To test that hypothesis, we evaluated the median 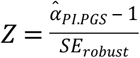 across the iterations. Second, across the iterations, we evaluated the median of the 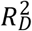 and the median difference in lifespan, analogous to **section 2**.**2**.**6**.

#### 2.2.9 Subjective life expectancy, self-rated health, and economic variables

We tested whether the 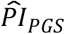 based on the Cox model (3) was associated with subjective life expectancy and self-rated health (**Supplementary Table 5**). Subjective life expectancy is defined as the ratio between respondents’ self-reported probability of surviving to a specific age divided by the life table prediction adjusted for age and sex. Because subjective life expectancy is normally distributed but left censored on 0, we performed both OLS and Tobit regressions on that outcome. Self-rated health was measured on a five-point Likert scale with the following categories: 1. Excellent; 2. Very good; 3. Good; 4. Fair; and 5. Poor. Thus, we estimated an ordinal logit regression. Both models controlled for the same covariates as our preferred model (3), and we clustered the standard errors at the household level.

We also tested the 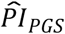 based on the Cox model (3) for association with the following 11 preregistered variables of relevance to economic research (**Supplementary Table 6**)^c^: (a) whether health limits the ability to work; (b) the self-reported probability of having a work-limiting health problem in the next 10 years; (c) the self-reported probability of working full-time after age 65; (d) plans to continue paid work in retirement; (e) concern about having enough retirement income; (f) planned retirement age (defined as planned retirement year minus birth year); (g) retirement satisfaction; (h) expectation of total retirement wealth; the percentage of waves covered by (i) life insurance or (j) by long-term care insurance; and (k) financial planning horizon. Depending on the distribution of each outcome, we applied either OLS, logit, or ordinal logit regression, with model (3) covariates and clustered standard errors.

## 3 Results

### 3.1 Results from the univariate survival analysis

With the 27 polygenic scores we constructed, we first performed univariate survival analyses of respondent, maternal, and paternal survival. The results are reported in **Supplementary Table 2** and **Supplementary Figures 3–5**. When survival was stratified by the top versus lower nine deciles of the score distribution, we found that 18 polygenic scores could significantly discriminate survival in either the respondents or their parents (at *P* < 0.05), and eight were Bonferroni-significant corrected for 27 traits. With respect to respondent survival, the following polygenic scores had a strong and significant influence (defined here as a >1 year difference in the median lifespan): (a) Alzheimer’s disease (1.2 y), (b) any ischemic stroke (1.3 y), (c) any stroke (1.6 y), (d) BMI (1.3 y), (d) cigarettes per day (2.2 y), (e) educational attainment (1.3 y), (f) prostate cancer (1.2 y), and (g) type 2 diabetes (1.6 y). As for the parents, the score for parental lifespan had the strongest influence (4 and 6 y in the mothers and fathers, respectively).

All univariate associations with respondent and parental survival were in the expected direction, except the association between respondent survival and the score for prostate cancer. (That score was not associated with parental survival.) Therefore, we performed an ad hoc robustness check to ensure that this score was associated with the likelihood of reporting a cancer diagnosis, and this association indeed was in the expected direction (*P* = 0.0007). Considered on its own, prostate cancer has an overall high survival rate as long as it is detected before metastasizing (Noone et al. 2018). We speculate that the genetic risk of prostate cancer may lead to certain health benefits if diagnosed early, such as more frequent doctor checkups or changes in lifestyle, which could explain the unexpected direction of effect; however, we emphasize that replication is necessary.

### 3.2 Results from the Cox proportional hazards regression

We report a selection of the multivariate survival analysis results in **Table 3** and the complete results in **Supplementary Table 3**. In our preferred model (3), we identified associations with the polygenic scores for Alzheimer’s disease 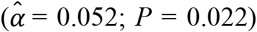, atrial fibrillation 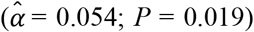, cigarettes per day 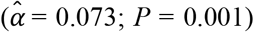, height 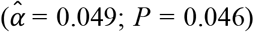, type 2 diabetes 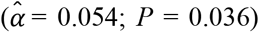, and parental lifespan 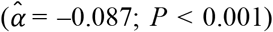. All estimated effects were in the anticipated direction. Importantly, we could not detect a violation of the PH assumption (*P* of the global *χ*^2^ = 0.404), and the model attained a satisfactory fit (e.g., a Cox and Snell *R*^2^ of 0.23). The 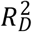 of the 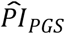 was 0.039 (95% CI = 0.028–0.052), while the 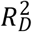 of the 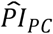 and 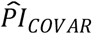 were 0.006 (0.002–0.011) and 0.113 (0.095–0.133), respectively. Thus, the polygenic scores explained substantially more of the variation on the log hazard scale than the genetic PCs, but it explained only about a third of the variation that the other model covariates did.

The only difference between model (2) and our preferred model (3) is the score for parental lifespan. That particular score was added separately because it may capture the influence of other scores, as suggested by the genetic correlations (**Supplementary Table 1** and **Supplementary Figure 2**). However, doing so did not notably influence the parameter estimates of the others, and the set of significant scores was the same between models (2) and (3). Notably, the effect of the score for parental lifespan in model (3) was the largest effect estimated for any of the polygenic scores across all four models 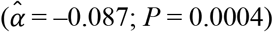.

Model (4) was our most extensively adjusted model; it additionally included the two health measures subjective life expectancy and self-rated health, various lifestyle factors (such as smoking and drinking), observed parental lifespan, and 11 indicators for categories of medical diagnoses, such as “ever diagnosed with cancer (or malignant tumor except skin cancer)”. Unfortunately, medical diagnoses were available only as binary indicators. Nevertheless, our primary interest is in model (3), which can be evaluated early in life. As could be expected by including directly observed health variables, model (4) drastically improved the model fit over both the null model and model (3) (both *P* ∼ 0). However, model (4) also strongly violated the PH assumption (*P* of the global *χ*^2^ = 0.00003). Reassuringly, the estimates of the polygenic scores, which we were particularly interested in, appeared highly stable across all four model specifications.

### 3.3 Results from the prognostic index analysis

With the Cox coefficients, we computed PIs for three sets of regressors: (i) the polygenic scores (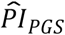), (ii) the genetic PCs (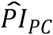), and (iii) the covariates (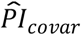). We first evaluated the relative influence of the three PIs within each model using the Royston & Sauerbrei 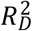 (**Supplementary Table 3**). Across the four models, the proportion of variation explained on the log hazard scale by the 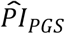 was between 0.03 and 0.041. The 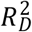 of that index was stable across the four models, which suggested that the polygenic scores explained a non-negligible proportion of variation even when we adjusted for socioeconomic variables, observable health risks, and other potential confounders.

Next, we used the PIs based on models (3) and (4) to stratify Kaplan-Meier survival functions, first using only 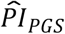 (**Table 4** and **Figure 1**). 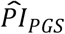 based on model (3) could distinguish a 3.5-year difference in median lifespan in comparison (a) between the top decile (*N* = 927) and the lower nine deciles (*N* = 8,345) and a 4.4-year difference in comparison (b) between the top decile and the bottom decile (*N* = 928). As expected, the 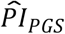 based on model (4) could distinguish somewhat less: 2.9 and 4.1 years in comparisons (a) and (b), respectively. The *P* values of the log-rank tests between the groups were all less than 7.91×10^−13^. Thus, the 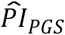 could distinguish a greater difference in median lifespan than any of the scores could on their own in the univariate analysis, even when based on the most extensively adjusted model (4).

Thereafter, we stratified survival using the three PIs simultaneously (**Table 5** and **Figure 2**). Here, we performed only comparison (a) (see **Methods**). With respect to the PIs based on model (3), respondents’ median lifespan was 3.1 years shorter in the top decile of 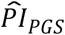 (*N* = 736) than in the lower nine deciles of all three PIs (*N* = 6,674) and was similar to the shorter median lifespan in the top decile of 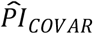 (3.9 y; *N* = 723). The few individuals who were in the top decile of both the 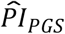 and the 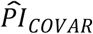 (*N* = 97) had an 8.1-year shorter median lifespan. The log-rank test was highly significant (*P* = 2.78×10^−75^).

As could be expected, in the analogous analysis based on model (4), the capacity of the 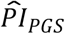 was somewhat reduced. Compared to the lower nine deciles of all three PIs (*N* = 6,605), the median lifespan was 2.4 years shorter in the top decile of the 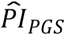 (*N* = 694), 10.7 years shorter for the 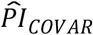 (*N* = 696), and an astonishing 14.8 years shorter for respondents in the top decile of both the 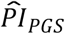 and the 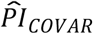 (*N* = 110). Respondents in the top decile of all three PIs had an 18.5-year shorter median lifespan, but we caution that this estimate is very noisy given the low number of observations in this cell (*N* = 11). Again, the log-rank test was highly significant (*P* = 3.59×10^−271^). Overall, adding health variables and medical conditions drastically increased the discriminatory capacity of the 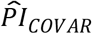, while it only slightly reduced the capacity of the 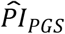, suggesting that the polygenic scores were able to add information above and beyond the inclusion of these intermediate variables.

#### 3.3.1 Benchmark to conventional actuarial risk factors

We benchmarked how well the 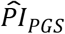 could distinguish median lifespan in comparison to conventional actuarial risk factors (**Supplementary Table 4** and **Supplementary Figure 6**). In that capacity, the 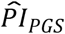 was comparable to sex (2.8 and 3.2 y in our study sample and the full sample of European HRS respondents, respectively), former smoker (2.5 and 3.4 y), and ever diagnosed with diabetes or high blood sugar (1.7 and 3.6 y). The 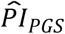 distinguished a greater difference than several conventional risk factors, including the top decile of years of schooling (corresponding to more than 16 years of schooling; 1.3 and 2 y), ever diagnosed with cancer (or malignant tumor except skin cancer; 1.2 and 1.7 y), and ever diagnosed with heart conditions (such as heart attack, coronary heart disease, angina, congestive heart failure, or other heart problems; 0.8 and 0.7 y). The 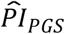 distinguished a smaller difference than the top decile of BMI (corresponding to BMI > 38.6; 4.4 and 5.3 y), current smoker (9.9 and 11.4 y), and ever diagnosed with chronic lung disease (except asthma, such as chronic bronchitis or emphysema; 4.3 and 4.3 y). Notably, these comparisons show that the ability of polygenic scores to classify individuals into groups of different mortality risks is similar to or better than that of several conventional actuarial risk factors when the polygenic scores are combined into our preferred genetic predictor, with the major difference that the prognostic index can be evaluated at a young age.

#### 3.3.2 Results of the cross-validation

We performed a cross-validation of model (3) to examine whether evaluating the prognostic indices in the full study sample could have introduced overfitting. Across 1,000 iterations, the median *Z* statistic of the test of the calibration slope was only marginally significant (*P* = 0.045). Similarly, the median 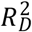 of the 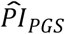 was 0.023 (instead of 0.039). The difference in median lifespan for comparisons (a) and (b) attenuated from 3.5 to 2.7 years and from 4.4 to 3.4 years, respectively. These attenuated differences in median survival fell within the confidence intervals of the main estimates and remained strongly significant. Thus, the cross-validation suggested that the ability to distinguish lifespan was somewhat overestimated when evaluated in the full sample, but reassuringly, our conclusions remain supported by the cross-validation.

#### 3.3.3 Results from the analysis of subjective life expectancy, self-rated health, and economic variables

We investigated whether the polygenic scores were associated with subjective life expectancy and self-rated health. The results are reported in **Supplementary Table 5**. Our genetic predictor of interest was the 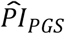 based on our preferred model (3), which had been standardized. For subjective life expectancy, the OLS and Tobit estimates were virtually identical, so we discuss only the Tobit results. The coefficient of the 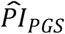 was estimated to be –0.052 (*SE* = 0.008; *P* = 8.03×10^−11^). As expected, greater genetic risk was associated with reporting an expectation of a shorter lifespan. However, the effect was small compared to, say, being female (–2.047; *SE* = 0.523), living in the western part of the United States (0.143; *SE* = 0.030), or being disabled (–0.298; *SE* = 0.101).

Next, we performed an ordinal logit regression of self-rated health (a higher value represents poorer health). In alignment with subjective life expectancy, we found that greater genetic risk was associated with reporting poorer health (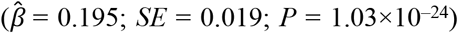 = 0.195; *SE* = 0.019; *P* = 1.03×10^−24^). Thus, assuming a proportional influence across the response categories, the odds ratio for being in a higher category was 1.215 per standard deviation of the 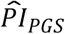. The effect was similar to that of being female (OR = 1.213) but much smaller than that of being disabled (OR = 8.315). Overall, these results indicate that genetic risks had indeed manifested and been observed via the respondent’s health.

Finally, we investigated whether the polygenic scores were associated with 11 economic variables. The results are reported in **Supplementary Table 6**. Our genetic predictor of interest, the 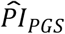 based on model (3), was significantly associated in the expected direction with five of the variables (*P* < 0.05) and Bonferroni-significant with three of the variables (*P* < 0.0045). That is, a standard deviation difference in the 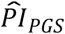 was associated with a higher probability of having or expecting a work-limiting health problem (OR = 1.202; *P* = 1.08×10^− 13^ and 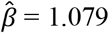; *P* = 8.66×10^−4^, respectively), less retirement satisfaction^d^ (OR = 1.132; *P* = 1.08×10^−6^), fewer waves of long-term care insurance coverage (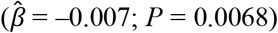; *P* = 0.0068), and a shorter financial planning horizon^e^ (OR = 0.953; *P* = 0.016). However, the estimated effect sizes were relatively small, which could be expected based on the currently limited signal in the polygenic scores. For example, per standard deviation, the 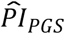 was associated with reporting a 1-percentage-point greater probability of having a work-limiting health problem in the next 10 years and with 2.2 months shorter long-term care insurance coverage. Nonetheless, the effects were comparable with those of, say, years of schooling, which was also associated with these five outcomes. Across the five outcomes, the difference compared to years of schooling was the greatest for long-term care insurance, where a standard deviation increase in years of schooling was associated with 10 months longer coverage.

We could not detect that greater genetic risk was associated with life insurance 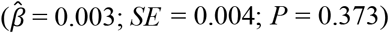. The estimated association corresponds to approximately 1 month longer life insurance coverage. In our study sample, we had only 14.5% statistical power to detect an effect of that size, which suggests that the association should be considered small. In contrast, we had 77.2% power to detect the aforementioned association with long-term care insurance, even though the difference in magnitude was minor. Nevertheless, our results show that polygenic scores for common diseases and mortality risks can already capture differences in economic outcomes, including insurance coverage.

## 4 Discussion

We investigated how well a broad set of polygenic scores for common diseases and mortality risks could distinguish differences in lifespan, and we benchmarked their performance in this regard to conventional actuarial risk factors. Our main finding is that polygenic scores have a joint capacity to classify people into groups of different mortality risk that is non-negligible, and this capacity is comparable to that of some conventional risk factors, including sex, former smoking, and years of schooling. Moreover, we found that the classification was even stronger than that of some of the diagnosed medical conditions that have been ascertained in the HRS, including ever diagnosed with cancer. Because we performed this comparison and adjusted more extensively for observable confounders, our results extend the literature on this topic. Importantly, the polygenic scores could explain a nontrivial part of the variation even in our most extensively adjusted model. We emphasize that our results represent only a lower bound of the predictive accuracy that polygenic scores will reach once larger GWAS become available (Daetwyler, Villanueva, and Woolliams 2008; de Vlaming et al. 2017).

Given these results, it is reasonable to expect that commercial interest in offering genetic tests for disease, mortality risks, and longevity to consumers is bound to increase further. Similarly, our results imply that polygenic scores already contain information that could be valuable to the insurance industry (e.g., in underwriting) if insurers were able to obtain genetic data from applicants and customers. Depending on whether genetic health information will remain legally or voluntarily exempt from underwriting, adverse selection will be more or less likely to occur as more people acquire knowledge of their genetic risks.

We found a significant but small negative association between genetic liability and long-term care insurance. This association may become stronger as the signal in polygenic scores increases. A conceivable mechanism for the association could be that elderly individuals who have observed a decline in their health (partly due to their unobserved genetic variants) have chosen not to purchase this insurance or have chosen to let it lapse, as they may expect not to reach an age that will require long-term care. The probability of requiring assistance with activities of daily living becomes more substantial after age 85 (Dionne 2013), and we found that individuals in the top decile of genetic risk reached just about that age, at the median. The associations of our preferred genetic predictor with subjective life expectancy and self-rated health support the idea that the respondents had indeed partly observed unobserved genetic risks. Thus, our results may imply that long-term care insurance is subject to weak adverse selection. However, an alternative explanation could be the high premium markups reported for this particular insurance type (Dionne 2013; Glenzer and Achou 2019), which could have rendered the product unattractive for people who consider themselves to be at risk of dying early or too expensive for people with work-limiting health problems.

On the contrary, we did not find that our preferred genetic predictor was associated with life insurance coverage, though this association may also become stronger in the near future. However, a null finding aligns with previous studies that have found little evidence of adverse selection in that market (Sijbrands, Tornij, and Homsma 2009; Harris and Yelowitz 2014; McCarthy and Mitchell 2010). Additionally, life insurance is typically purchased at middle age, before many heritable medical conditions have had time to manifest (Nabholz and Rechfeld 2017). Thus, the difference between life insurance and long-term care insurance could be explained by the latter more often being purchased at an older age (Cornell et al. 2016). Nonetheless, the genetic data we studied here were most likely unobserved by the HRS participants since most waves of data were collected prior to the advent of consumer genetic testing for disease. In addition, most of the polygenic scores we generated are not yet part of genetic health reports offered to consumers. Thus, we should perhaps not expect that extensive self-selection has already occurred.

Next, our results imply that consumers will at some point have knowledge of genetic risks that they may be incentivized to disclose when purchasing insurance products tied to their survival. This would apply, e.g., in the market for “enhanced annuities” (that is, life annuities underwritten not only with demographic information but also with medical information). Applicants at greater risk could potentially benefit from lower premiums (or higher benefits) if their genetics risks were underwritten (Veyssiere et al. 2017; Steinorth 2012). It has been reported that standard-rate life annuities may be actuarially unfair in the United States because premiums are determined using low-mortality assumptions to counter potential adverse selection (as it is assumed that this product is bought primarily by the healthiest and wealthiest) (Brown and Scahill 2010). In such a market, the possibility of revealing individual risks could benefit people with a reduced life expectancy who consider the standard rate expensive. At the same time, some experts argue that enhanced annuities could crowd out the standard rate product, and genetic testing may potentially exacerbate that development (Steinorth 2012). Overall, we consider further investigation of consumer behavior under conditions of private knowledge of genetic risks an interesting avenue of future research.

We think that as the accuracy of genetic predictors matures and more consumers acquire private knowledge of their genetic risks, genetic health information may eventually have to be treated just like any other kind of medical information that can currently be requested by insurance providers (Strohmenger and Wambach 2000). There are many scholars who argue that providers are already entrusted with handling very private and sensitive information, such as medical journals and tests, and that there should be no reason to expect them to handle genetic information with any less prudence (Rothstein 2004). However, some of them consider it unethical to charge more for genetic factors that are outside of the control of the carrier, though a counterargument is that many conventional risks are also outside the control of the affected. Additionally, there is a real risk that insurance coverage may be denied to those who receive a burdening outcome in the “genetic lottery”, which results in greater need for solidarity (Prince 2019). Importantly, it could be detrimental to public health if people avoid genetic testing for medical or research purposes out of fear of discrimination (Keogh et al. 2017).

Thus, strong arguments can be made that actuarial discrimination of any kind based on genetic factors should be restricted (Rothstein 2018; Newson et al. 2018). Additionally, we emphasize that genetic risk factors influencing common diseases are not deterministic, and some act via non-biological pathways, such as lifestyle choices, that are yet undetermined or that may be modifiable (Khera et al. 2019). These complications question the ethicality of classifying mortality risks based on genetic test results. Overall, we encourage policymakers, the insurance industry, and other stakeholders to monitor this development closely and to have a scientifically informed discussion about the potential consequences of determining premiums or rejections based on genetic information on the one hand and adverse selection on the other.

Our results should be considered in light of a few limitations. First, it is likely that mortality selection has led to an underestimation of the current performance of polygenic scores. This would have foremost affected the scores for conditions that manifest at younger age. For example, in contrast to our expectations, we did not find an association with the score for coronary artery disease, which is a major cause of NCDs (Lloyd-Jones et al. 2006). Another important limitation, which is endemic to the GWAS literature (Clyde 2019; Mills and Rahal 2019), is that we did not study individuals of non-European ancestry. Thus, we do not know whether our findings generalize to other ancestries (Martin et al. 2019, 2017). Unfortunately, it will take many years before we can thoroughly answer that question. Lastly, we acknowledge that the medical conditions ascertained in the HRS are based on self-reports and lack specificity, and it could be that their true impact on survival is understated in our analyses. Future studies with access to richer medical data will have to determine more precisely how much information polygenic scores can add above and beyond various biomarkers for disease and already acquired medical conditions.

Our results bear on the ongoing debate on whether and how to regulate the commercialization of genetic health information (Allyse et al. 2018; Van Hellemondt, Hendriks, and Breuning 2011; Howard and Borry 2012; Kricka et al. 2011; Moscarello et al. 2019; Badalato, Kalokairinou, and Borry 2017). Many companies sell genetic tests with limited guidance and lengthy disclaimers, leaving their customers puzzled or confused (Nelson, Bowen, and Fullerton 2019; Wang et al. 2018). For example, it could be financially detrimental for the dependents of a life insurance policy if the policy holder voluntarily terminates their policy out of a false belief of low genetic risk. Therefore, we agree with others who call for more extensive consumer protection in this market (Schleit, Naylor, and Hisama 2019). Overall, more research is needed to determine how vulnerable types of consumers, for example, those with particularly high genetic risk or those with weak genetic literacy, react after exploring their DNA for health information. In particular, genetic tests with low accuracy are easy to misinterpret (Schleit, Naylor, and Hisama 2019). Thus, for now, we think that advertisement of disease and longevity predictions is ethically questionable at many levels and should be done only with great care, though the appropriateness of such services of course depends on how they are marketed and how the results are presented. However, at this moment, many genetic testing services hide behind extensive disclaimers to void them of responsibility, which could be considered a questionable practice (Schleit, Naylor, and Hisama 2019).

Therefore, as a policy recommendation, we encourage regulatory authorities to consider prognostic genetic testing for disease and longevity to be a form of genetic counseling. As such, we would consider it reasonable to limit the practice to licensed or accredited institutions, be they public or private. However, standards for genetic counseling are still maturing globally (Ormond et al. 2018; Abacan et al. 2019), and until regulatory measures are taken and an industry standard has been established, it is likely that appropriate consumer protection will lag behind technological developments. At the same time, since many people appear eager to purchase genetic health information, we consider it undesirable to completely restrict an individual persons’ right to explore their DNA, with or without the assistance of a certified counselor. Additionally, the borderless nature of genetic testing, where consumers can send their genetic data to services located in different jurisdictions, makes it practically impossible to effectively regulate this market at only the national level. Thus, appropriate regulation of consumer genetics will require international agreements to be effective.

In conclusion, the estimates presented here clearly show the relevance of polygenic scores in the context of insurance. However, much research is required before it can be determined which polygenic scores may potentially meet the criteria for evidence-based underwriting and before accurate and fair premiums could be developed. Ultimately, policymakers and regulatory agencies will have to strike a difficult balance between keeping private insurance fair and viable on the one hand while ensuring satisfactory consumer protection against genetic discrimination and privacy violations on the other. In the meantime, depending on the jurisdiction, we see a tangible risk of both genetic discrimination and informational advantage on the consumer side, which could lead to adverse selection.

## Data Availability

Data are available in research repositories. Sources are described in detail in the manuscript.

https://www.ncbi.nlm.nih.gov/projects/gap/cgi-bin/study.cgi?study_id=phs000428.v2.p2

http://hrsonline.isr.umich.edu/index.php?p=shoavail&iyear=X7&_ga=2.116438468.379878193.1585556093-979669872.1584616185

## 5 Author contributions

This study was conceived by R.K.L. and designed by R.K.L together with P.D.K. P.D.K oversaw and supervised the study. The preregistered analysis plan was written by R.K.L. with critical input by P.D.K. R.K.L performed the analyses. R.K.L. and P.D.K together interpreted the results. R.K.L wrote the manuscript with critical input from P.D.K. R.K.L. produced the tables and figures.

## 6 Acknowledgments

The research was approved by the Research Ethics Review Board (RERB) at the School of Business and Economics (SBE) of Vrije Universiteit Amsterdam (20180927.1.pkr730). We gratefully acknowledge Netspar for financially supporting this research project with a topicality grant (BD2018.01) and thank Bas Werker and Anja de Waegenaere for their valuable feedback on the research questions, study design, and draft manuscript. We are thankful to Casper Burik for constructing the genetic PCs and Aysu Okbay for a providing a meta-analysis of educational attainment that excluded the Health and Retirement Study (HRS). This work was carried out on the Dutch national e-infrastructure (grant 17148) with support of the SURF Cooperative. Philipp Koellinger was financially supported by an ERC Consolidator Grant (647648 EdGe). HRS is supported by the National Institute on Aging (NIA U01AG009740). The genotyping was funded separately by the National Institute on Aging (RC2 AG036495, RC4 AG039029). Genotyping was conducted by the NIH Center for Inherited Disease Research (CIDR) at Johns Hopkins University in Baltimore, Maryland. Genotyping quality control and final preparation of the data were performed by the Genetics Coordinating Center at the University of Washington in Seattle, Washington. Genotype data can be accessed via the database of Genotypes and Phenotypes (dbGaP, http://www.ncbi.nlm.nih.gov/gap, accession number phs000428.v2.p2). Researchers who wish to link genetic data with other HRS measures that are not in dbGaP, such as educational attainment, must apply to the HRS for permission. See the HRS website (http://hrsonline.isr.umich.edu/gwas) for details.

## Footnotes

a The analysis plan is available at https://osf.io/qzx6p/

b In this study, we adhere to the definition of European ancestry that is standard in genetic epidemiology, which distinguishes “Hispanic/Latin American” (Mills and Rahal 2019).

c We also preregistered that we would study private health-insurance coverage. However, we found little variation in health-insurance coverage in the HRS, and thus, we dropped this particular analysis from power considerations.

d Retirement satisfaction was coded as “1. very; 2. moderately; or 3. not at all”.

e Financial planning horizon was coded as “1. next few months; 2. next year; 3. next few years; 4. next 5-10 years; or 5. longer than 10 years”.

